# Skin Residual Bilirubin Volume (SRBV): A Physiologically Informed Framework for Transcutaneous Bilirubin Interpretation in Neonates

**DOI:** 10.64898/2026.03.03.26347511

**Authors:** HO Amadi

## Abstract

**Background:** Neonatal jaundice management increasingly relies on transcutaneous bilirubinometry (TcB), yet discrepancies with serum bilirubin (TSB) have limited its clinical reliability. This study introduces Skin Residual Bilirubin Volume (SRBV) as a physiologically grounded framework to enhance TcB interpretation.

**Objective:** To evaluate SRBV as an explanation for TcB–TSB discordance and assess whether incorporating SRBV improves the interpretability and reliability of TcB measurements during diagnosis, phototherapy, and recovery.

**Methods:** TcB readings (MBj20) were calibrated against laboratory TSB in non-jaundiced neonates (TSB <3 mg/dL). Neonates undergoing phototherapy were monitored using paired TcB measurements before and after treatment breaks (TBL-out and TBL-return). TSB was measured before treatment, at mid-treatment, and prior to discharge. Patterns of TcB–TSB disparity and an observed reproducible ‘Recovery Value Flip (RVP)’ phenomenon were analysed.

**Results:** Across 102 neonates, TBL consistently equalled or exceeded TSB, supporting the additive SRBV model. Early in phototherapy, TBL-return > TBL-out, indicating persistent cutaneous bilirubin. A reproducible RVP occurred mid-treatment, after which TBL-return < TBL-out coincided with sustained bilirubin decline. Fractional SRBV contribution increased with baseline bilirubin and persisted into recovery, demonstrating dynamic, patient-specific cutaneous bilirubin retention.

**Conclusion:** SRBV provides a biologically plausible explanation for TcB–TSB discordance and dynamic TcB behaviour. Incorporating SRBV into TcB interpretation enables physiologically informed monitoring, improving safety and reliability in laboratory-limited neonatal settings.

**Significance Statement:** Transcutaneous bilirubinometry is widely used but limited by disagreement with serum bilirubin. This study introduces SRBV as a physiological explanation for TcB variability and proposes an SRBV-adjusted framework that transforms TcB measurements into actionable, non-invasive clinical guidance.

## 1. Introduction

Neonatal jaundice, the clinical manifestation of elevated bilirubin in newborns, is one of the most common conditions encountered in postnatal care worldwide. Physiologic jaundice occurs in the majority of term and preterm infants during the first week of life, affecting up to 60–85 % of neonates and contributing substantially to hospital readmission and healthcare utilisation globally. Severe or untreated hyperbilirubinemia can lead to acute bilirubin encephalopathy, long-term neurodevelopmental sequelae, and even death, particularly in low-resource settings where timely diagnosis and treatment are challenging [1].

Accurate measurement of bilirubin levels is essential for identifying neonates at risk and guiding phototherapy decisions. The gold standard for bilirubin quantification remains total serum bilirubin (TSB) obtained via laboratory analysis of blood samples. However, in many low- and middle-income countries (LMICs) and remote or resource-limited health facilities, access to laboratory services is often unavailable, delayed, or prohibitively expensive. This limitation reduces timely initiation of therapy and increases the risk of bilirubin toxicity [2].

To address these challenges, transcutaneous bilirubinometry (TcB) was developed as a non-invasive, point-of-care alternative that estimates bilirubin by measuring skin reflectance. TcB devices have been shown to correlate with TSB in many settings and are widely used to screen for neonatal hyperbilirubinemia. Moreover, universal TcB screening has been demonstrated to be more effective than visual inspection alone in identifying neonates requiring intervention, reducing unnecessary invasive sampling, and aiding discharge planning [3].

Despite these advantages, several studies have identified important limitations in the reliability and accuracy of TcB measurements. Early investigations reported that although TcB correlates with TSB, it cannot consistently predict absolute serum bilirubin values, particularly at higher concentrations, and is therefore limited as a standalone diagnostic tool. TcB has generally been recommended as a screening rather than a definitive diagnostic modality, with careful selection of decision thresholds to minimise false-negative results [4].

Phototherapy—the mainstay of jaundice treatment—introduces further complexity to TcB interpretation. Some studies have found that TcB measurements during or soon after phototherapy may be less accurate due to changes in skin reflectance and bilirubin distribution, with reports of only moderate correlation with TSB during these phases. Others have shown that even when measured after phototherapy, TcB may underestimate serum levels or demonstrate poor agreement, underscoring the need for laboratory confirmation when available [5–7].

Factors such as skin pigmentation, measurement site, gestational age, and device type have also been shown to influence TcB accuracy and agreement with TSB, limiting universal application without correction or contextual understanding [6].

These persistent inconsistencies in agreement between transcutaneous bilirubin levels (TBL) and TSB have limited TcB’s adoption for diagnostic decision-making, treatment monitoring, and discharge planning in settings without laboratory access. This gap highlights the need for approaches that not only improve measurement accuracy but also contextualise TcB readings within the underlying bilirubin physiology—especially in environments where laboratory tests are scarce.

This study addresses a fundamental question: why does transcutaneous bilirubinometry behave inconsistently, and how can it be made more reliable without additional equipment?

## 2. Conceptual Framework: TSB, TBL, and SRBV

### 2.1 Defining Skin Residual Bilirubin Volume (SRBV)

TSB represents the absolute concentration of bilirubin within the bloodstream, measured invasively via blood sampling. In contrast, TBL is obtained using optical techniques that interrogate bilirubin within and beyond the intravascular compartment (Figure 1).

**Figure 1.**
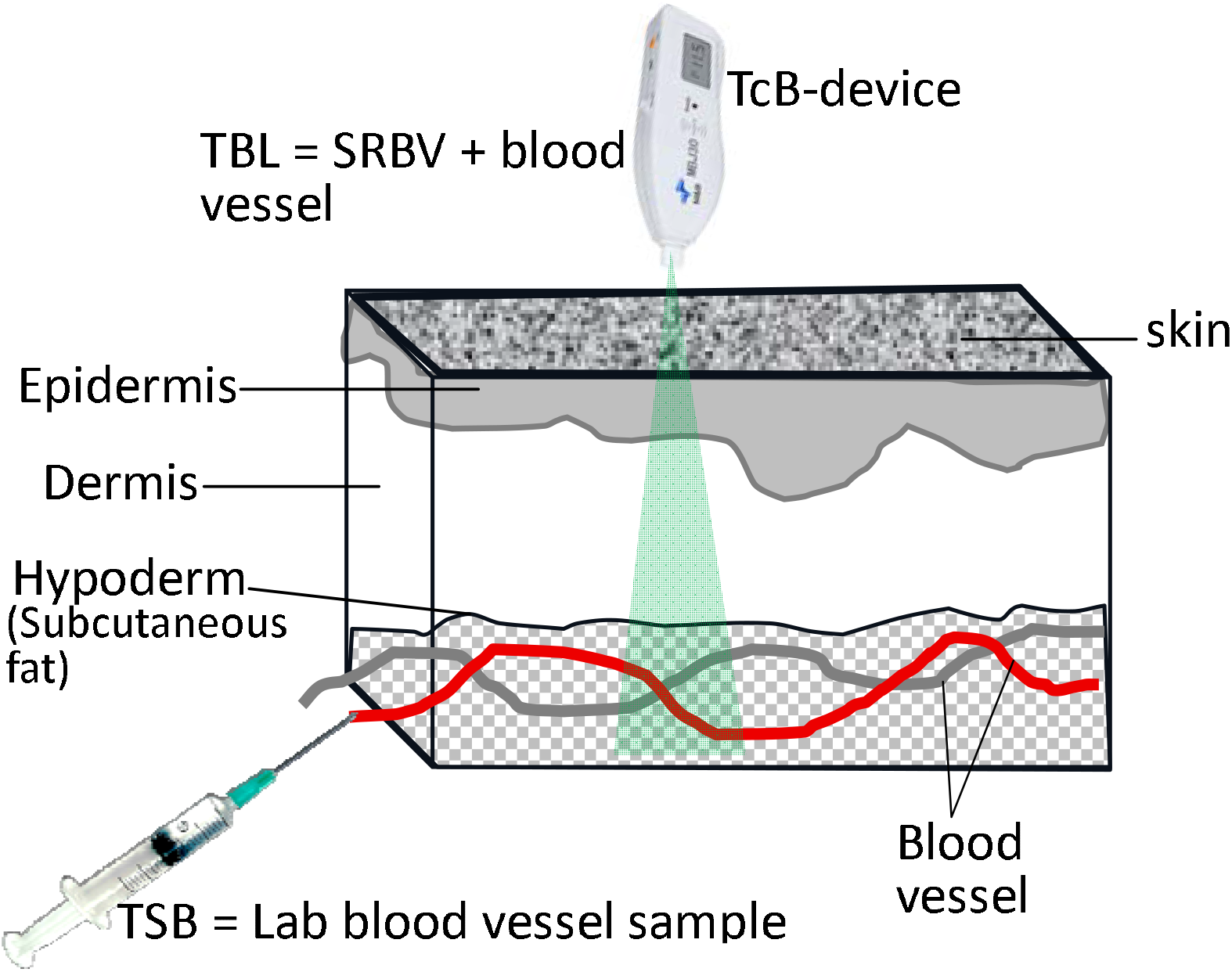
TSB, TBL, SRBV concept

Figure 1:

We propose that TcB devices inherently measure:

- Intravascular bilirubin (corresponding to TSB), plus
- Residual bilirubin retained in the skin and subcutaneous tissues, termed Skin Residual Bilirubin Volume (SRBV)

Thus:

TBL ≈ TSB + SRBV

In neonates with very low bilirubin concentrations (TSB <3 mg/dL), bilirubin remains intravascular and SRBV ≈ 0. Under these conditions, TBL closely approximates TSB.

At higher bilirubin levels, bilirubin diffuses into the skin, generating a non-zero SRBV and causing TBL to exceed TSB.

### 2.2 Hypothetical Pathophysiology

Bilirubin extravasation into the skin produces visible jaundice and reflects a dynamic equilibrium between serum and tissue compartments. The threshold at which this occurs varies between patients, influenced by skin thickness, perfusion, maturity, and prior bilirubin exposure.

Consequently, identical TSB values may correspond to different SRBV magnitudes in different neonates, particularly during recovery following phototherapy. This variability provides a physiological basis for TcB–TSB discordance.

## 3. Materials and Methods

### 3.1 Study Design and Setting

This was a prospective, multicentre observational study conducted across 10 grassroots mission hospital facilities in the southeastern region of Nigeria as part of an ongoing task-shift implementation programme for neonatal jaundice management in laboratory-limited settings [8]. The study focused on evaluating the physiological basis and clinical interpretability of transcutaneous bilirubin (TcB) measurements during neonatal jaundice treatment.

### 3.2 Transcutaneous Bilirubinometer Calibration

The MBj20 transcutaneous bilirubinometer (Beijing M&B Electronic Instruments, Beijing, China) was calibrated locally at each participating facility prior to clinical deployment. Calibration was performed using paired transcutaneous bilirubin level (TBL) and laboratory-based total serum bilirubin (TSB) measurements obtained from neonates without clinical jaundice (TSB <3 mg/dL).

For calibration, TcB measurements were obtained from the forehead concurrently with venous blood sampling for laboratory TSB analysis. Paired values were used to derive a centre-specific calibration coefficient, which was applied to the TcB device before routine clinical use at that facility. This approach was intended to minimise device and site-related measurement variability and to ensure consistency across centres.

### 3.3 Study Objectives

The study was designed to address the following objectives:

1. To establish whether TBL measurements are consistently equal to or greater than TSB across patients and bilirubin severity categories.
2. To characterise the magnitude of skin residual bilirubin volume (SRBV) across clinically relevant bilirubin ranges.
3. To examine SRBV behaviour near treatment completion, particularly when TBL values fall below 11 mg/dL.
4. To determine whether SRBV exhibits time-dependent behaviour during phototherapy and recovery.

### 3.4 Treatment Protocol

Neonates diagnosed with jaundice received phototherapy using the solar-powered PoliteUltraLumen phototherapy device, which was deployed uniformly across all participating mission hospitals [9]. Phototherapy was delivered in 3-hour continuous treatment sessions, separated by 20-minute breaks to allow feeding and rehydration.

### 3.5 Monitoring and Data Collection

During each treatment break, TcB measurements were obtained using the calibrated MBj20 device at a standardised anatomical site (forehead) and recorded as:

- TBL-out: measured immediately at cessation of a phototherapy session
- TBL-return: measured immediately before resumption of the subsequent session

Laboratory blood samples for TSB quantification were collected before initiation of treatment, at least once during mid-treatment, and prior to discharge, in accordance with the clinical protocol at each centre. These paired TcB and TSB measurements were used to assess TcB–TSB disparity and infer SRBV dynamics during treatment and recovery.

### 3.6 Ethical Considerations

Ethical approval for the study was obtained from the Ethics Committee of the Catholic Diocese of Nsukka, Nigeria. Written informed consent was obtained from the parent or caregiver of each neonate prior to enrolment. All procedures were conducted in accordance with applicable ethical standards and local clinical governance requirements.

## 4. Results

### 4.1 Study Population and Consistency of TBL ≥ TSB

A total of 102 neonates were recruited across six of the ten participating grassroots mission hospitals (Table 1), covering a wide spectrum of jaundice severity with baseline transcutaneous bilirubin levels ranging from 6.8 to 30.4 mg/dL. Due to financial constraints, three centres were unable to perform laboratory TSB measurements, and among these, only one facility completed full TSB sampling as specified in the protocol, while the remaining centres obtained partial TSB data.

**Table 1.** Recruitment by centre, baseline transcutaneous bilirubin characteristics, treatment-phase bilirubin dynamics, and manifestation of the Recovery Value Flip (RVP).

| TBL-TSB behavioural values |  |  |  |  |  |  |  |
| --- | --- | --- | --- | --- | --- | --- | --- |
| Hospital | Total Patient | Ave Post-natal Age (days) | Ave pre-treatment TBL | TBL range | TBL drop consistency (mg/dL/hr) |  | RVP manifest: from >9mg/dL |
|  |  |  |  |  | Ave TBL decline rate | end-treatment: TBL≥TSB count |  |
| **BSH Nsukka** | 23 | 5.74 | 14.03 | 10—28 | 0.27 | all | 91% |
| **SPH Abakaliki** | 23 | 3.83 | 12.37 | 8.9—18.4 | 0.52 | NA | 100% |
| **MJH Okigwe** | 10 | 5.7 | 12.29 | 10.4—14.9 | 0.23 | NA | 100% |
| **IHH Urualla** | 20 | 2.7 | 8.97 | 6.8—14.3 | 0.21 | NA | 71% |
| **MCH Enugu** | 13 | 3.83 | 16.33 | 10.7—23.5 | 0.18 | NA | 92% |
| **IHMH Nkpor** | 13 | 3.71 | 15.22 | 8.3—30.4 | 0.18 | all | 100% |

(BSH – Bishop Shanahan Hospital, MCH – Mother of Christ Hospital, IHMH – Immaculate Heart of Mary Hospital, SPH – St Patrick Hospital, MJH – Mbanjo Joint Hospital, IHH – Immaculate Heart Hospital)

Across all bilirubin categories in which paired measurements were available, transcutaneous bilirubin levels (TBL) were consistently equal to or greater than corresponding total serum bilirubin (TSB) values. This relationship was observed irrespective of centre, baseline bilirubin level, or treatment stage. In several patients, the magnitude of TBL–TSB disparity increased discretely with rising bilirubin levels. Early TBL measurements below 3 mg/dL demonstrated negligible TBL–TSB difference, whereas higher baseline values were associated with measurable disparities, reflecting inter-individual variability.

Table 1 summarises recruitment by centre, baseline bilirubin ranges, average pre-treatment TBL values, observed TBL decline rates during treatment, and the frequency with which end-treatment TBL values remained equal to or greater than TSB.

### 4.2 Recovery Value Flip (RVP) During Phototherapy

Serial TBL measurements obtained during phototherapy revealed a reproducible dynamic pattern among neonates with baseline TBL values greater than 9 mg/dL. Early in treatment, TBL measured immediately at the end of a phototherapy session (TBL-out) was consistently lower than TBL measured immediately before the subsequent session (TBL-return). This pattern was observed across centres and treatment sessions.

After several phototherapy cycles, a Recovery Value Flip (RVP) was observed, characterised by a reversal of this relationship such that TBL-return became consistently lower than TBL-out (Table 1). Following the occurrence of RVP, TBL declined steadily until treatment completion. The RVP phenomenon was clearly identifiable in 92% of patients with adequate serial TcB data and was observed across a range of baseline bilirubin severities.

Figure 2 illustrates representative TBL trajectories demonstrating the RVP phenomenon and the subsequent monotonic decline in bilirubin levels until discharge.

**Figure 2.**
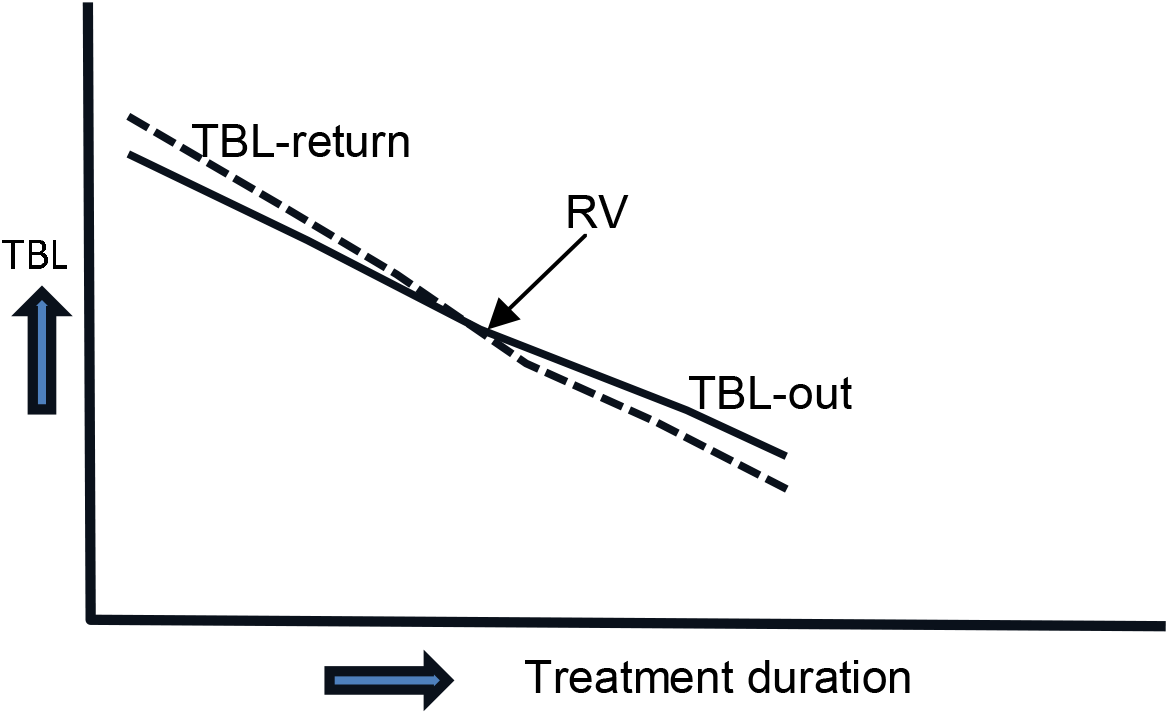
Representative transcutaneous bilirubin trajectories illustrating the Recovery Value Flip (RVP) phenomenon and subsequent decline during phototherapy.

### 4.3 Fractional Contribution of SRBV to Measured TBL

The estimated fractional contribution of skin residual bilirubin volume (SRBV) to measured TBL is summarised in Table 2, stratified by bilirubin severity category and treatment stage. SRBV was expressed as a proportion of TBL and averaged within clinically relevant bilirubin ranges.

**Table 2.** Estimated average fractional contribution of skin residual bilirubin volume (SRBV) to measured transcutaneous bilirubin (TBL) before treatment and during recovery.

| Hospital/GROUP | %SRBV in TBL |  |  |  |  |  |
| --- | --- | --- | --- | --- | --- | --- |
|  | Pre-treatment |  |  | End-treatment |  |  |
|  | <9<br>mg/dL | 9-11.5<br>mg/dL | 11.6-15<br>mg/dL | <9<br>mg/dL | 9-11.5<br>mg/dL | 11.6-15<br>mg/dL |
| **BSH Nsukka** | NA | 23% | 9% | 17% | 27% | 24% |
| **MCH Enugu** | NA | 11% | 12% | NA | NA | NA |
| **IHMH Nkpor** | NA | NA | NA | 21% | 11% | 28% |
| **Average** |  | 17% | 11% | 19% | 19% | 26% |
| **SD** |  | 0.08 | 0.02 | 0.03 | 0.11 | 0.03 |

(BSH – Bishop Shanahan Hospital, MCH – Mother of Christ Hospital, IHMH – Immaculate Heart of Mary Hospital)

At pre-treatment, measurable SRBV contribution was observed primarily in neonates with mild to moderate jaundice. The average SRBV fraction was 17% (SD 0.08) in the 9–11.5 mg/dL group and 11% (SD 0.02) in the 11.6–15 mg/dL group. In neonates with TBL <9 mg/dL, SRBV was not detectable, and insufficient data were available to reliably estimate pre-treatment SRBV in neonates with TBL >15 mg/dL.

During the recovery stage, SRBV contribution was detectable across a broader range of bilirubin values. Neonates with TBL <9 mg/dL demonstrated a mean SRBV fraction of 19% (SD 0.03). In the 9–11.5 mg/dL and 11.6–15 mg/dL groups, mean SRBV fractions increased to 19% (SD 0.11) and 26% (SD 0.03), respectively. These fractional estimates were observed consistently across centres where sufficient paired data were available.

## 5. Discussion

### 5.1 Interpreting Transcutaneous Bilirubinometry Through SRBV

This study reframes transcutaneous bilirubinometry from a flawed surrogate of serum bilirubin into a composite physiological signal. The persistent observation that transcutaneous bilirubin levels (TBL) were equal to or greater than total serum bilirubin (TSB) across bilirubin strata supports the conceptual model in which TcB reflects the sum of intravascular bilirubin and skin residual bilirubin volume (SRBV). Apparent TcB “overestimation,” particularly at higher bilirubin levels and during phototherapy, therefore, reflects physiological bilirubin persistence within the skin rather than device inaccuracy.

Importantly, this relationship was dynamic rather than static, evolving across treatment phases. The observed variability in TBL–TSB disparity between patients is consistent with inter-individual differences in bilirubin extravasation, cutaneous deposition, and clearance, reinforcing the need for interpretation frameworks that extend beyond isolated numeric thresholds.

### 5.2 Clinical Meaning of the Recovery Value Flip (RVP)

Serial TcB monitoring during phototherapy revealed a reproducible phenomenon termed the Recovery Value Flip (RVP). Early in treatment, TBL measured before therapy resumption (TBL-return) exceeded values measured immediately after phototherapy cessation (TBL-out), consistent with ongoing bilirubin redistribution into the skin during treatment breaks. As treatment progressed, a distinct inflection point was reached at which TBL-return became lower than TBL-out, signalling depletion of the cutaneous bilirubin reservoir and dominance of sustained bilirubin clearance.

The consistency of the RVP across centres and patients suggests that it represents a clinically meaningful physiological milestone rather than random measurement variability. The RVP therefore provides a non-invasive marker of treatment adequacy that is particularly valuable in settings where laboratory confirmation is unavailable.

### 5.3 SRBV as a Unifying Explanation for TcB–TSB Discordance

By explicitly accounting for SRBV, this study provides a physiological explanation for the long-recognised discordance between TcB and TSB reported across devices, populations, and treatment contexts. Rather than indicating poor device performance, TcB variability appears to encode information about bilirubin compartmentalisation and redistribution.

The graded SRBV fractions observed across bilirubin strata and treatment stages (Table 2) further support this interpretation. SRBV emerged with increasing bilirubin burden, persisted during recovery, and declined only after sustained treatment response, confirming TcB as a biologically interpretable measurement rather than a direct serum analogue.

### 5.4 SRBV-Adjusted TcB Interpretation as a Proposed Standard of Care

Based on these findings, we propose an SRBV-adjusted TcB interpretation framework for use in laboratory-limited neonatal care settings (Figure 3). This approach departs from reliance on isolated TcB values and instead emphasises contextual, trend-based interpretation incorporating treatment phase and SRBV dynamics.

**Figure 3.**
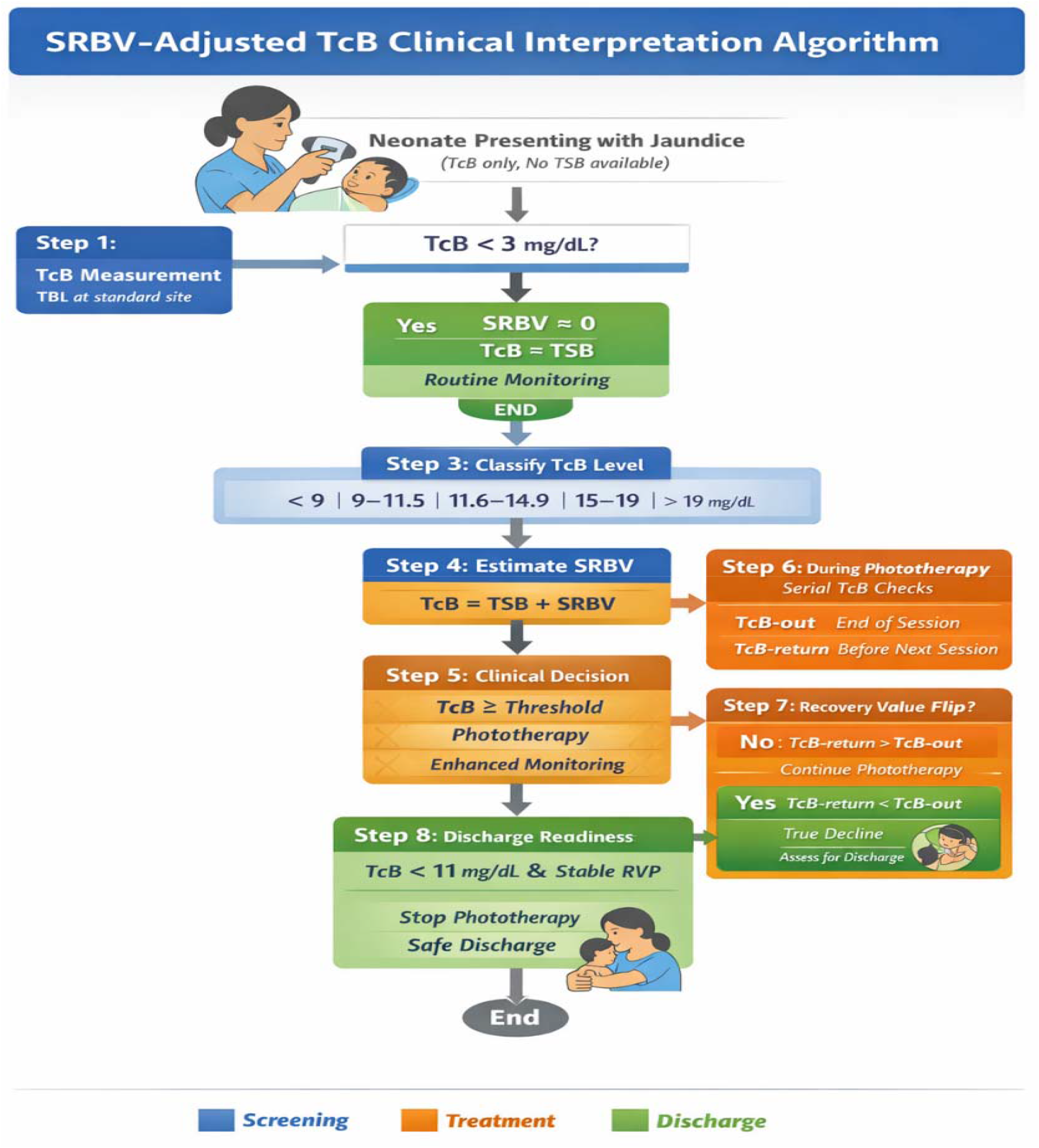
SRBV-adjusted transcutaneous bilirubin interpretation algorithm

Under this proposed standard of care:

1. TcB values <3 mg/dL reliably approximate serum bilirubin, reflecting negligible SRBV.
2. TcB values ≥3 mg/dL are assumed to include a variable SRBV component and should not be interpreted as direct serum equivalents.
3. Paired TcB measurements during phototherapy breaks inform compartment dominance:
  - TBL-return > TBL-out indicates SRBV persistence.
  - Observation of the RVP (TBL-return < TBL-out) signals depletion of cutaneous bilirubin.
4. Post-RVP TcB trends are prioritised over absolute values for treatment continuation, cessation, and discharge decisions.

Within this framework, the RVP functions as a non-invasive physiological marker of recovery, enabling safer TcB-guided care without laboratory confirmation.

### 5.5 Implications for Neonatal Care in Laboratory-Limited Settings

As outlined in the Introduction, neonatal jaundice continues to cause preventable morbidity and mortality in settings where access to laboratory-based TSB testing is limited or delayed. The SRBV-adjusted TcB interpretation framework directly addresses this gap by extending the clinical reliability of TcB in contexts where its use is unavoidable.

By accounting for skin-related bilirubin persistence, this approach expands TcB utility beyond screening to treatment monitoring and discharge planning, including during and after phototherapy—scenarios traditionally considered unreliable for TcB interpretation. The framework supports task-shifting to nurses, midwives, and community providers using a structured, physiology-informed approach and aligns naturally with decentralised and solar-powered phototherapy programmes. At a system level, it has the potential to reduce unnecessary blood sampling, avoid overtreatment, and shorten hospital stays.

### 5.6 Positioning SRBV Within Existing Guidelines

Current neonatal jaundice guidelines appropriately caution against standalone TcB use at higher bilirubin levels and during phototherapy. The SRBV framework provides a physiological rationale for these cautions while offering a pathway to overcome them in constrained settings. By explicitly accounting for cutaneous bilirubin persistence and redistribution, SRBV-adjusted TcB interpretation may serve as an interim or complementary standard of care where laboratory confirmation is unavailable.

### 5.7 Limitations and Future Directions

This study has several limitations. Findings were derived from a limited number of TcB devices, and device-specific optical characteristics may influence SRBV behaviour. Measurements were obtained predominantly from the forehead, and regional variation in skin properties may affect SRBV expression at alternative sites. SRBV was quantified indirectly from systematic TcB–TSB disparity rather than measured as an absolute tissue parameter, constraining mechanistic precision. In addition, partial laboratory data availability limited assessment of population-level modifiers such as gestational age and skin pigmentation.

Further multicentre validation across diverse populations, devices, and clinical environments is required. Nevertheless, the consistency of observed TcB–TSB relationships and treatment-phase dynamics establishes SRBV as a clinically meaningful construct and provides a rational foundation for the proposed interpretation framework.

## 6. Conclusion

This study introduces skin residual bilirubin volume (SRBV) as a unifying physiological construct explaining the systematic disparity between transcutaneous and serum bilirubin measurements, particularly at higher bilirubin levels and during phototherapy. By recognising TcB as a composite of intravascular and skin-deposited bilirubin, the SRBV framework reframes TcB variability from a limitation into clinically interpretable information. Integration of SRBV into a structured interpretation algorithm extends the role of TcB beyond screening to treatment monitoring and discharge decision-making in laboratory-limited settings. While further validation is required, SRBV represents a pragmatic and scalable advance aligned with the realities of neonatal care delivery in resource-constrained environments.

Significance Statement: Transcutaneous bilirubinometry is widely used in low-resource settings but has been limited by poor agreement with serum bilirubin at clinically relevant levels. This study introduces skin residual bilirubin volume (SRBV) as a physiological explanation for this disparity and proposes an SRBV-adjusted interpretation framework that transforms TcB variability into actionable clinical information. The approach advances safer, non-invasive jaundice management in settings where reliance on TcB is a necessity rather than a choice.

## Data Availability

All data produced in the present study are available upon reasonable request to the authors

## 7. Data availability

De-identified data generated during this study will be deposited in the Imperial College London institutional repository and made publicly available upon publication.

## 10. Acknowledgements

I wish to acknowledge the kind contributions of some paediatricians, neonatologists and others during the design of this study, particularly, Dr Emmanuel Asogwa, Professor Martin Meremikwu, Professor Obot Antia-Obong, Dr Sara Ango, Dr Leo-Stanley Muoneke, And Professor Jacob J Udo. The medical missions of Professor H Amadi in Nigeria is anchored by Neonatal Concerns for Africa (www.neonatalconcerns.org)—a collaboration of the Bioengineering Department of Imperial College London, and in part, supported by the Hornchurch Baptist Church, Essex England, United Kingdom (https://www.hornchurchbaptist.org.uk). I acknowledge all the kind collaborative support of the nine Catholic Bishops and Dioceses whose grassroots hospitals participated in this project across the southeast of Nigeria. I wish to particularly thank their Lordships Most Rev Dr Jonas Benson Okoye (the Catholic Bishop of Nnewi) and Most Rev Professor Godfrey Igwebuike Onah (the Catholic Bishop of Nsukka) for their unwavering encouragement. I acknowledge the prayers and encouragements from the Rev Mother Mauren Akabogu, the Mother Superior General of the sisters of Immaculate Heart of Mary Mother of Christ congregation and Rev Sr Dr Nkiruka Okafor, the congregational Health CEO, for her boundless support throughout this project. Worthy of mention is the tireless diocesan neonatal care teams at our grassroots “Jaundiroom centres” in Nsukka, Nkpor, Enugu, Okigwe, Urualla, and Abakaliki. I exceptionally praise the hard work of ward-manager Mr Raphael Okechukwu Odo and Rev Fr Osmond Anike and their jaundiroom-team members at Nsukka for producing the most comprehensive data utilized in this analysis. The basic version of ChatGPT was applied during the final editing process of this manuscript.

